# Changes in consumer purchasing patterns at New York City chain restaurants following adoption of the sodium warning icon rule, 2015-2017

**DOI:** 10.1101/2022.08.22.22279098

**Authors:** Divya Prasad, John P. Jasek, Amaka V. Anekwe, Christine Dominianni, Tamar Adjoian Mezzacca, Julia S. Sisti, Shannon M. Farley, Kimberly Kessler

## Abstract

In 2016, New York City (NYC) began enforcing a sodium warning regulation at chain restaurants, requiring placement of an icon next to any menu item containing ≥2,300 mg sodium. As shifts in consumer purchases are a potential outcome of menu labeling, we investigated whether high-sodium purchases from NYC chains changed following policy implementation. Consumer purchases, using receipts for verification, were assessed at 2 full-service (FSR) and 2 quick-service (QSR) chain restaurants in NYC and Yonkers, NY, which did not have sodium menu labeling, in 2015 (baseline) and 2017 (follow-up). Difference-in-difference regression models, adjusted for demographic and location co-variates, tested whether the proportion of NYC respondents purchasing high-sodium item(s) (containing ≥2,300 mg sodium) or whether mean sodium content of purchases changed, in FSR or QSR. Relative to Yonkers, the proportion of NYC respondents purchasing 1 or more high-sodium items did not significantly differ from baseline to follow-up at FSR (difference-in-difference: -4.6%, p=0.364) or QSR (difference-in-difference: -8.9%, p=0.196). Among NYC FSR respondents, mean sodium purchased significantly declined compared to Yonkers (difference-in-difference: -524 mg, p=0.012); no changes in mean sodium were observed among QSR participants (difference-in-difference: 258 mg, p=0.185). While the reduction in mean sodium purchased among NYC FSR patrons after the sodium warning implementation is encouraging, there was not a corresponding reduction in the proportion of respondents purchasing high-sodium items. Further research that evaluates longer-term changes in the restaurant environment, including menu offerings and consumer purchases, following such policies is warranted.

## Introduction

In 2016, New York City (NYC) began enforcing a sodium warning regulation. This novel policy requires chain restaurants (those with 15 or more locations nationally) to display warning icons next to high-sodium menu items and post a statement explaining the icon and associated health risks [1] (S1 Fig) [2]. Sodium in the restaurant environment is problematic, given that restaurant food constitutes more than a quarter of the sodium consumed by US adults [3], the sodium content of restaurant offerings is often high [4-5], and between 2003-2016, the amount of sodium consumed by adults from full-service restaurant (FSR) meals increased and the amount from fast-food, or quick-service restaurant (QSR) meals, remained high [6]. Average sodium consumption among both US [7] and NYC [8] adults far exceeds the 2,300 milligram (mg) Chronic Disease Risk Reduction level established by the National Academies of Sciences [9] and adopted by the Dietary Guidelines for Americans [7], and lowering sodium intake is associated with reductions in hypertension and cardiovascular disease risk [10].

Prior to the passage of NYC’s sodium warning regulation, numerical sodium content labeling on menus was implemented in three other US jurisdictions: Philadelphia, PA [11], King County, WA [12] and Pierce County, WA [13]. All three efforts also involved displaying other nutrition information, such as calories and fat, alongside menu items. Though Philadelphia has since adopted sodium warning icon labeling like NYC’s [14], a consumer purchase evaluation of its previous policy found that the sodium content of purchases at a subset of FSR chains subject to Philadelphia’s labeling policy was lower than that at control chains outside of Philadelphia [9].

We assessed consumer purchases at a subset of FSR and QSR chains in NYC and in Yonkers, NY, a control city, before and after enforcement of NYC’s sodium warning regulation, with the objective of determining whether high-sodium items, sodium, and calories in purchases changed in NYC.

## Methods

### Study sample

Baseline data were collected at chain restaurants in NYC and Yonkers between October 2015 and January 2016. Yonkers was selected as a control city for its similar demographic profile and existing calorie labeling requirement at chain restaurants, which was also in effect in NYC. Although the sodium warning regulation went into effect in December 2015, enforcement did not begin until June 2016 and none of the chains included in the final analysis implemented the regulation during the baseline data collection period. Follow-up data were collected in both cities between April-June 2017.

Rationale and methods behind selection of restaurant chains and locations have been previously described [15]. Briefly, FSR and QSR chains with the greatest number of NYC locations and with at least one location in 3 out of the 5 NYC boroughs and Yonkers were identified as: Applebee’s, IHOP and TGI Friday’s (FSR) and Burger King, McDonald’s, Popeyes and Subway (QSR). From these restaurant chains, at least one FSR and one QSR location in each NYC borough was included. The final analytic sample of restaurants consisted of 2 FSR chains: IHOP and TGI Friday’s, and 2 QSR chains: Popeyes and Subway. Baseline data collection at Applebee’s was terminated early due to this chain’s advance implementation of the sodium warning regulation. Burger King and McDonald’s were excluded from the analytic sample because few or no items met criteria for the sodium warning icon.

At FSR, surveys were conducted at 9 NYC and 1 Yonkers IHOP locations, and at 4 NYC and 1 Yonkers TGI Friday’s locations. At QSR, surveys were conducted at 3 NYC and 1 Yonkers Popeyes locations, and at 6 NYC (5 at follow-up due to closure of 1 location) and 2 Yonkers Subway locations.

### Measures

#### Survey

Surveys were conducted by trained interviewers between 5-9 pm at FSR locations on weekdays and weekends, and between 12-3 pm at QSR on weekdays. All adult patrons leaving locations were invited to participate in the survey for a $5 incentive. Patrons without an itemized receipt were ineligible. Participants were asked to identify items on their receipt that were purchased for their own consumption, whether and how they modified default menu items (e.g., extra cheese, side salad instead of fries), and whether and how often they refilled a beverage. The survey also collected respondents’ demographic characteristics, including self-reported age group, race/ethnicity, educational level and interviewer-observed gender. Collecting demographic data by observation is not a best practice at the NYC Department of Health and Mental Hygiene (DOHMH) and may have resulted in misclassification since gender cannot be accurately determined by observation.

#### Sodium and calories

Items listed on the receipt that the respondent verified as for their personal consumption were included. Sodium and calorie values for these items were obtained from MenuStat.org (MenuStat), a free, public database that publishes nutrition information from websites of the largest US restaurant chains. This study used nutrition information collected in January 2015 for items purchased at baseline and January 2017 for items purchased at follow-up. Nutrition information could not be obtained for all purchased items, as menu offerings can fluctuate throughout the year or by location, and as MenuStat does not include information for alcoholic beverages. Participants missing nutrition information for their entire purchase were excluded from analysis.

#### High-sodium items

Baseline purchases were coded as “high-sodium items” (S1-4 Tables) if they met criteria for displaying a warning icon – if the item itself had or could be customized to contain 2,300 mg of sodium or more, or if a combination meal was purchased that had the potential to contain at least 2,300 mg of sodium. Photos of menu boards and menu books were taken at a single NYC location for each chain during both data collection periods. If an item purchased at follow-up displayed a sodium warning icon in the restaurant’s follow-up menu photos, it was coded as a high-sodium item. If, at follow-up, a purchased item was not shown on the menu photos, it was coded using the same logic as baseline items.

The DOHMH’s IRB reviewed the study protocol and determined it to be exempt human subjects’ research.

### Additional exclusions

Prior to analysis, participants were excluded if their purchase could not be verified (e.g., items reported as being consumed did not match those listed on the receipt) or if only a beverage was purchased. Extreme outliers, defined as purchases exceeding 12,000 mg of sodium, were excluded. Daily purchases per location were also examined for unusual patterns; “outlier days,” where most purchases were solely for a side item or a beverage, were identified at 1 NYC Subway location, 1 Yonkers Subway location, and 1 Yonkers IHOP location, all at baseline. All purchases made on these dates at these locations were excluded. Figs 1-2 show the numbers of FSR and QSR participants excluded for each of these reasons.

**Fig 1.**
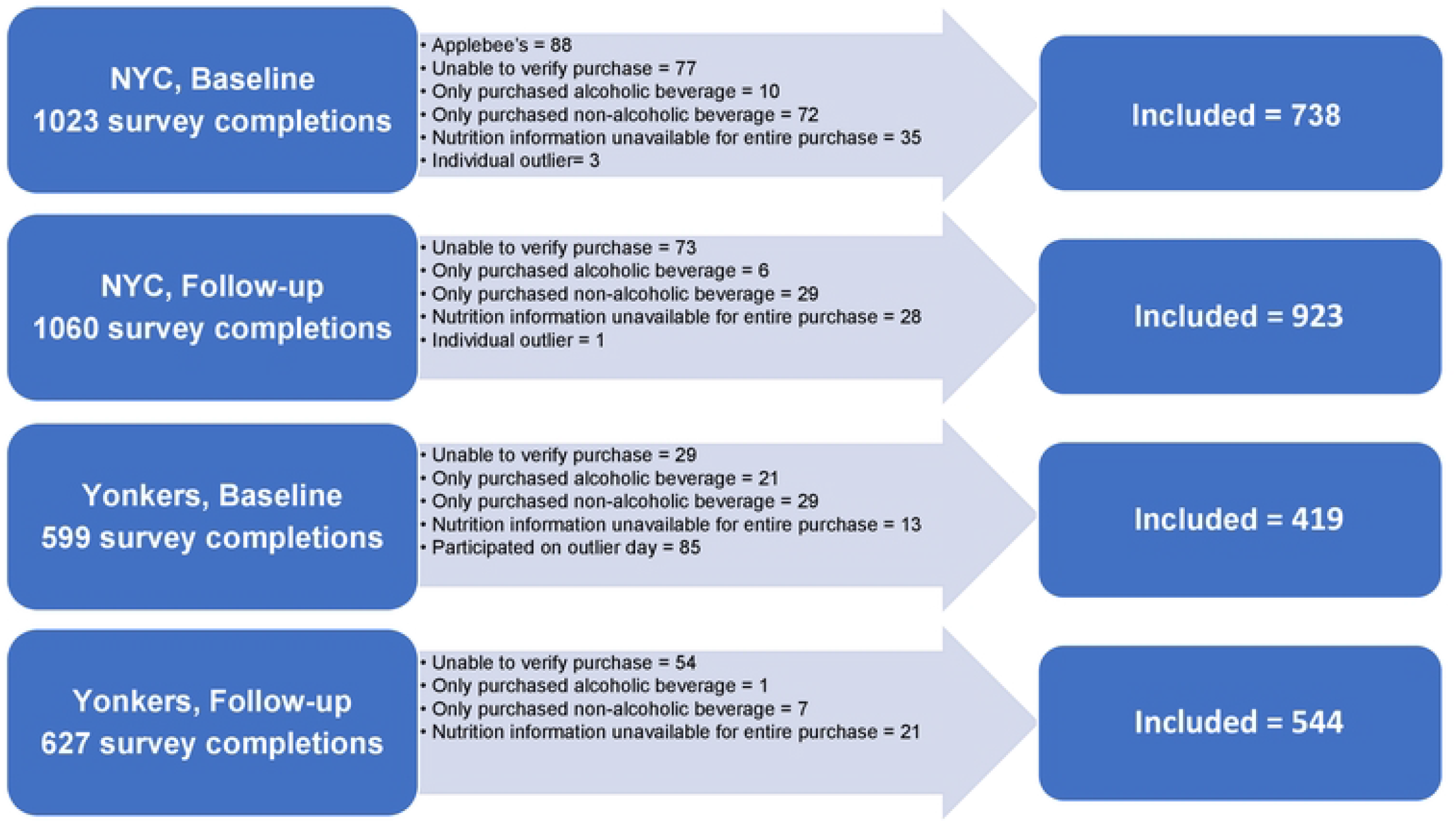
Participant Flow Chart, Full-Service Restaurants.

**Fig 2.**
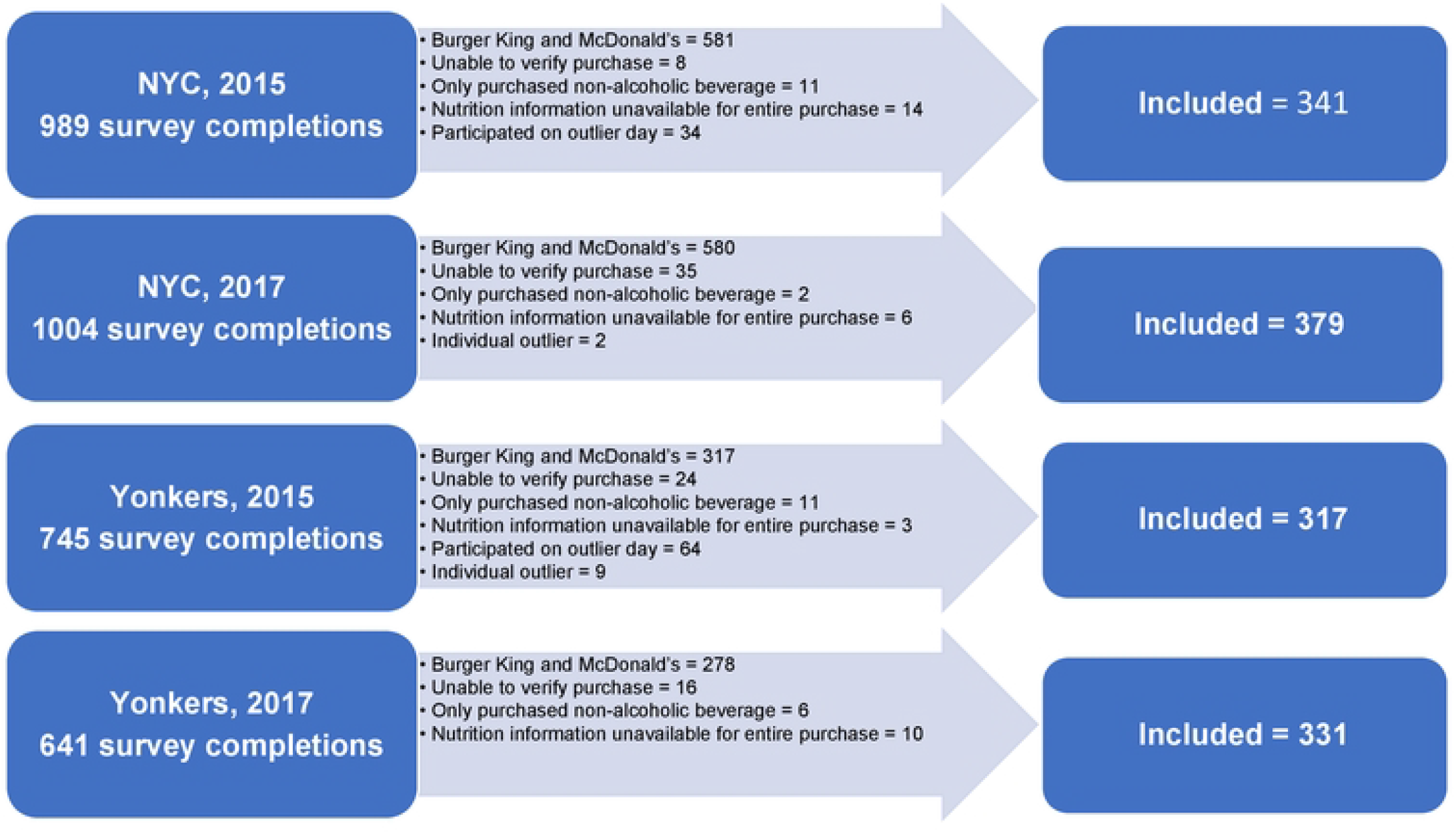
Participant Flow Chart, Quick-Service Restaurants.

### Statistical analysis

Sample characteristics were summarized and compared between cities and timepoints via chi-squared tests.

Analyzed outcomes included number of high sodium items purchased, and sodium and calorie content of purchases. Estimates and difference-in-differences between NYC and Yonkers from baseline to follow-up were assessed via mixed-effects regression models. Mean high-sodium items purchased were estimated by Poisson regression. The likelihood of purchasing at least 1 (both FSR and QSR) and at least 2 (FSR only) high-sodium items was assessed using logit regression. Purchases of 2 or more high-sodium items were extremely rare (among less than 1% of the sample) at QSR and therefore were not assessed. Mean sodium and calories purchased were estimated via linear regression models. To reduce skewness and meet model residual normality assumptions, sodium and calorie values were square-root-transformed to assess differences. However, for ease of interpretation, least-squared means estimates from models using non-transformed sodium and calories are reported here. Model estimates for both non-transformed and transformed sodium and calories are shown in S5 Table.

All models included fixed effects for restaurant chain, gender, age group, race/ethnicity and education, as these differed between cohorts, and random effects for restaurant street address.

Data were analyzed in SAS Enterprise Guide 7.1 (SAS Institute Inc., Cary, NC, USA).

## Results

### Participants

The final sample of respondents consisted of: at NYC FSR, 738 baseline, 923 follow-up; at Yonkers FSR, 419 baseline, 544 follow-up; at NYC QSR, 341 baseline, 379 follow-up; at Yonkers QSR, 317 baseline, 331 follow-up.

Restaurant chain, gender, age, race/ethnicity, and educational attainment of participants are summarized for each year and city in Tables 1 (FSR) and 2 (QSR). Sample distribution differed across most of these characteristics significantly between years within each city, and between cities within each year.

**Table 1:** Participant characteristics at baseline and follow-up, full-service restaurants.

|  | NYC |  |  | Yonkers |  |  | NYC vs Yonkers |  |
| --- | --- | --- | --- | --- | --- | --- | --- | --- |
|  | Baseline<br>n=738<br>n (%) | Follow-up<br>n=923<br>n (%) | p value | Baseline<br>n=419<br>n (%) | Follow-up<br>n=544<br>n (%) | p value | p, 2015 | p, 2017 |
| **Chain** |  |  |  |  |  |  |  |  |
| IHOP | 631 (85.5) | 633 (68.6) | **<.001** | 146 (34.8) | 495 (91.0) | **<.001** | **<.001** | **<.001** |
| TGI Friday's | 107 (14.5) | 290 (31.4) |  | 273 (65.2) | 49 (9.0) |  |  |  |
| **Gender** |  |  |  |  |  |  |  |  |
| Man | 385 (52.2) | 327 (35.4) | **<.001** | 229 (54.7) | 205 (37.7) | **<.001** | 0.416 | 0.236 |
| Woman | 353 (47.8) | 618 (64.6) |  | 190 (45.3) | 337 (62.0) |  |  |  |
| Other/Unknown | 0 (0.0) | 0 (0.0) |  | 0 (0.0) | 1 (0.2) |  |  |  |
| Missing | 0 (0.0) | 0 (0.0) |  | 0 (0.0) | 1 (0.2) |  |  |  |
| **Age (years)** |  |  |  |  |  |  |  |  |
| 18-24 | 181 (24.5) | 151 (16.4) | **0.004** | 79 (18.9) | 36 (6.6) | **<.001** | 0.133 | **<.001** |
| 25-34 | 194 (26.3) | 265 (28.7) |  | 117 (27.9) | 96 (17.6) |  |  |  |
| 35-44 | 199 (27.0) | 276 (29.9) |  | 115 (27.4) | 238 (43.8) |  |  |  |
| 45-64 | 142 (19.2) | 197 (21.3) |  | 87 (20.8) | 135 (24.8) |  |  |  |
| 65+ | 21 (2.9) | 32 (3.5) |  | 21 (5.0) | 39 (7.2) |  |  |  |
| Refused | 1 (0.1) | 2 (0.2) |  | 0 (0.0) | 0 (0.0) |  |  |  |
| **Race/Ethnicity** |  |  |  |  |  |  |  |  |
| Asian/Pacific Islander, non-Latino | 46 (6.2) | 18 (2.0) | **<.001** | 12 (2.9) | 6 (1.1) | 0.270 | **<.001** | **<.001** |
| Black, non-Latino | 241 (32.7) | 359 (38.9) |  | 169 (40.3) | 224 (41.2) |  |  |  |
| Latino | 264 (35.8) | 314 (34.0) |  | 100 (23.9) | 130 (23.9) |  |  |  |
| White, non-Latino | 148 (20.1) | 205 (22.2) |  | 123 (29.4) | 170 (31.3) |  |  |  |
| Other, non-Latino | 36 (4.9) | 22 (2.4) |  | 14 (3.3) | 11 (2.0) |  |  |  |
| Don't Know/Refused | 3 (0.4) | 5 (0.5) |  | 1 (0.2) | 3 (0.6) |  |  |  |
| **Education** |  |  |  |  |  |  |  |  |
| Less than HS | 38 (5.2) | 19 (2.1) | **<.001** | 17 (4.1) | 3 (0.6) | **<.001** | 0.238 | **<.001** |
| High School Grad | 223 (30.2) | 270 (29.3) |  | 147 (35.1) | 124 (22.8) |  |  |  |
| Some College | 268 (36.3) | 280 (30.3) |  | 153 (36.5) | 234 (43.0) |  |  |  |
| College Grad | 209 (28.3) | 353 (38.2) |  | 102 (24.3) | 182 (33.5) |  |  |  |
| Refused | 0 (0.0) | 1 (0.1) |  | 0 (0.0) | 1 (0.2) |  |  |  |

**Bolded** p values indicate statistically significant differences (p < 0.05)

**Table 2:**
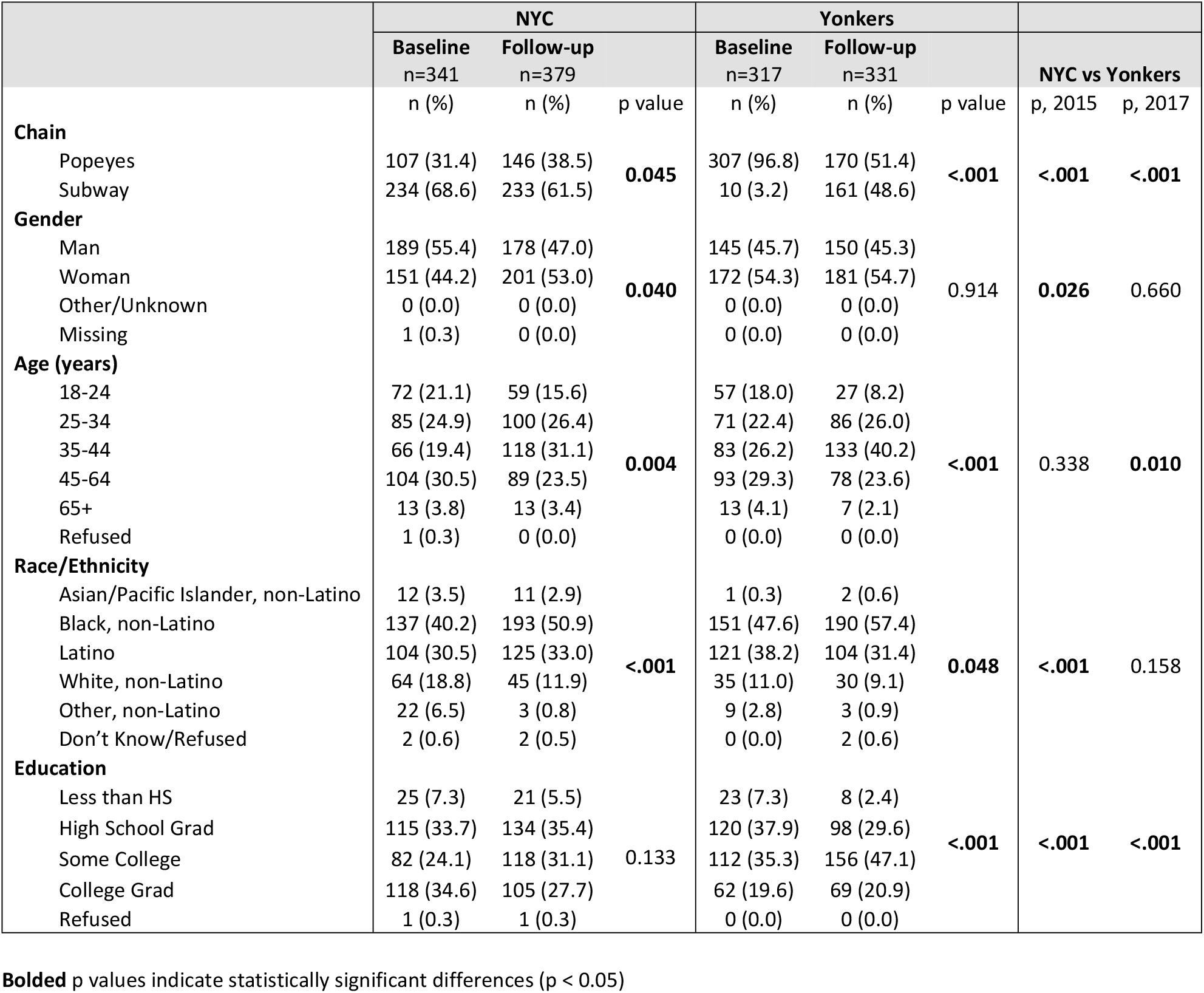
Participant characteristics at baseline and follow-up, quick-service restaurants.

### High-sodium item purchases (Table 3)

Among NYC FSR respondents, there was a significant decline from baseline to follow-up in the mean number of high-sodium items purchased (0.83 vs 0.65 items/participant, p<0.001) and the proportion of respondents purchasing at least 2 high-sodium items (15% vs 3%, p<0.001). The proportion of respondents purchasing at least 1 high-sodium item did not differ between baseline and follow-up (67% vs 65%, p=0.389).

**Table 3:** Purchases at full-service and quick-service restaurant chains, pre- (baseline) and post- (follow-up) implementation of the sodium warning icon in NYC.

|  | NYC vs<br>Yonkers | NYC (intervention) |  |  |  | Yonkers (control) |  |  |  |
| --- | --- | --- | --- | --- | --- | --- | --- | --- | --- |
|  | Difference<br>In<br>Difference* | Baseline |  | Follow-up |  | Baseline |  | Follow-up |  |
| **Full-Service Restaurants** |  | n=734 |  | n=915 |  | n=418 |  | n=538 |  |
| # High sodium items purchased per participant, mean (95% CI) | -0.13 | 0.83 | (0.74, 0.93) | **0.65<sup>a</sup>** | (0.58, 0.73) | 0.67 | (0.59, 0.76) | 0.62 | (0.53, 0.71) |
| Participants purchasing at least 1 high-sodium item, % (95% CI) | -4.6 | 67.4 | (61.1, 73.1) | 65.0 | (58.6, 70.8) | 59.3 | (51.5, 66.7) | 61.5 | (53.8, 68.7) |
| Participants purchasing at least 2 high-sodium items, % (95% CI) | -7.5 | 15.0 | (9.9, 22.0) | **3.1<sup>a</sup>** | (1.7, 5.4) | 6.1 | (3.6, 10.3) | **1.7<sup>b</sup>** | (0.6, 4.4) |
| **Quick-Service Restaurants** |  | n=338 |  | n=376 |  | n=317 |  | n=329 |  |
| # High sodium items purchased per participant, mean (95% CI) | -0.03 | 0.25 | (0.19, 0.34) | 0.27 | (0.20, 0.36) | 0.25 | (0.19, 0.34) | 0.30 | (0.22, 0.41) |
| Participants purchasing at least 1 high-sodium item, % (95% CI) | -8.9 | 26.8 | (18.2, 37.7) | 30.3 | (20.8, 41.9) | 24.6 | (16.3, 35.5) | **37.0<sup>b</sup>** | (25.7, 50.0) |

Models were adjusted for restaurant chain, gender, age group, education, and race/ethnicity, with random effects for restaurant location (street address)
Sample sizes for participants included in the models are shown. Due to missingness in fixed effects variables, 19 FSR and 8 QSR participants in the final analytic sample were not included in modeled estimates.
\* all p values for difference-in-differences were greater than 0.05
**Bolded** estimates indicate statistically significant differences between baseline and follow-up
<sup>a</sup> 2017 vs 2015 p < 0.001
<sup>b</sup> 2017 vs 2015 p < 0.05

Among Yonkers FSR respondents, there was a significant decline from baseline to follow-up in the proportion of respondents purchasing at least 2 high-sodium items (6% vs 2%, p=0.014). The mean number of high-sodium items purchased by Yonkers FSR respondents did not differ between baseline and follow-up (0.67 vs 0.62 items/participant, p=0.309), nor did the proportion of respondents purchasing at least 1 high-sodium item (59% vs 62%, p=0.598).

At FSR, there was no significant change from baseline to follow-up in NYC relative to Yonkers (i.e., difference-in-difference) in the mean number of high-sodium items purchased (p=0.120) or in the proportion of respondents purchasing at least 1 (p=0.364) or at least 2 (p=0.509) high-sodium items.

Among NYC QSR respondents, there was no significant difference between baseline and follow-up in the mean number of high-sodium items purchased (0.25 vs 0.27 items/participant, p=0.603), or in the proportion of participants purchasing at least one of these items (27% vs 30%, p=0.422).

Among Yonkers QSR participants, there was a significant increase from baseline to follow-up (25% vs 37%, p=0.027) in the proportion of participants purchasing at least one high-sodium item. The mean number of high-sodium items did not significantly differ between baseline and follow-up (0.25 vs 0.30 items/participant, p=0.112).

At QSR, there was no difference-in-difference between NYC and Yonkers in the mean number of high-sodium items purchased (p=0.463) or in the proportion of respondents purchasing at least 1 high-sodium item (p=0.196).

### Sodium and Calories

Mean sodium content of purchases declined significantly from baseline to follow-up at both NYC FSR (3,245 mg vs 2,279 mg, p <0.001) and Yonkers FSR (2,696 mg vs 2,254 mg, p=0.016). The decline in NYC was significantly greater than that in Yonkers (p=0.012). Similar patterns were present for the mean calorie content of purchases, with declines at both NYC (baseline: 1,524 kcal; follow-up: 1,055 kcal; p <0.001) and Yonkers (baseline: 1,259 kcal; follow-up: 1,009 kcal; p=0.004), and a significantly greater decline in NYC than in Yonkers (p=0.02). (Fig 3).

**Fig 3:**
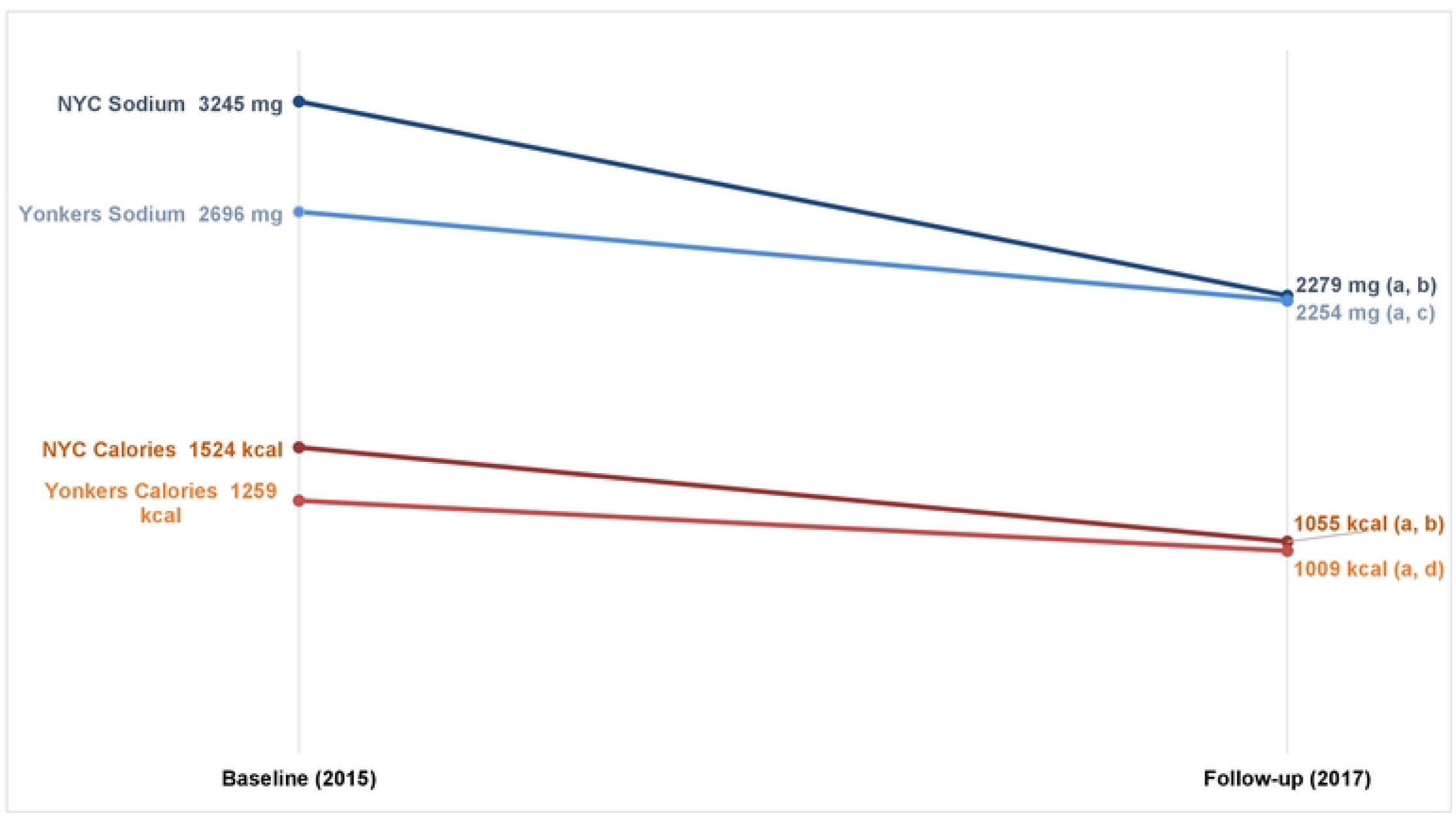
Mean sodium and calorie content* of purchases at FSR chains, pre- (Baseline, 2015) and post-(Follow-up, 2017) implementation of the sodium warning icon in NYC. *Sodium and calorie values were square-root-transformed during analysis to meet model normality assumptions and to estimate differences (p values), but least-squared means estimates from models using non-transformed variables are presented for ease of interpretation. Full model estimates are shown in S5 Table. a) NYC vs Yonkers Difference-in-Difference p value < 0.05; b) 2015 vs 2017 p value < 0.001; c) 2015 vs 2017 p value < 0.05; d) 2015 vs 2017 p value <0.01.

At QSR, mean sodium content of purchases did not significantly differ from baseline to follow-up in NYC (1,977 mg vs 1,777 mg, p=0.162) or in Yonkers (2,193 mg vs 1,735 mg, p=0.064), and there was no difference-in-difference between the two cities (p=0.570). Mean calories purchased declined from baseline to follow-up in both NYC (946 kcal vs 794 kcal, p=0.013) and in Yonkers (1,047 kcal vs 836 kcal, p=0.039), with no difference-in-difference between the two cities (p=0.768). (Fig 4).

**Fig. 4:**
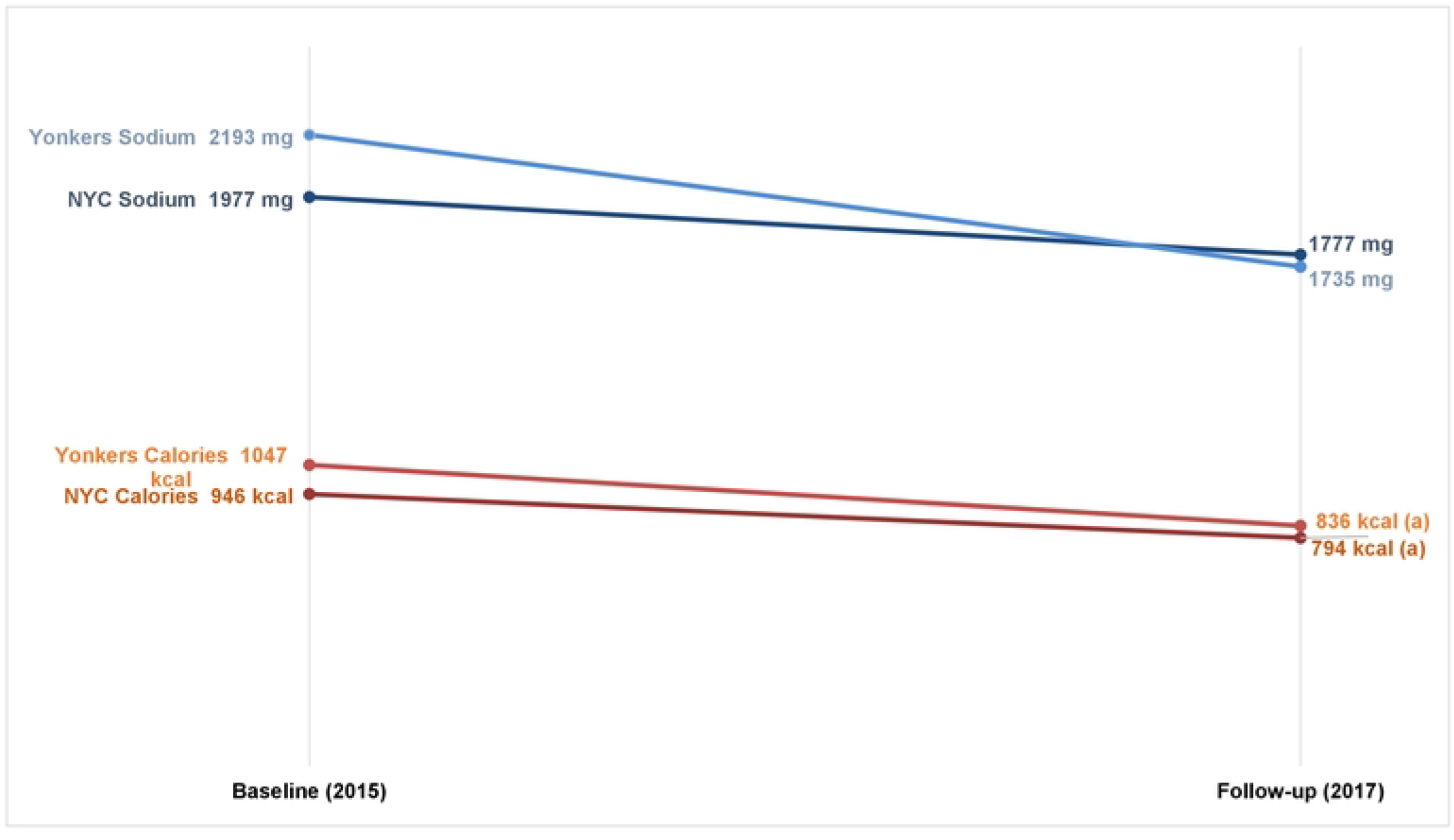
Mean sodium and calorie content* of purchases QSR chains, pre- (Baseline, 2015) and post-(Follow-up, 2017) implementation of the sodium warning icon in NYC. *Sodium and calorie values were square-root-transformed during analysis to meet model normality assumptions and to estimate differences (p values), but least-squared means estimates from models using non-transformed variables are presented for ease of interpretation. Full model estimates are shown in S5 Table. a) 2015 vs 2017 p value < 0.05

## Discussion

Here, we find evidence of changes in purchasing patterns at NYC FSR following implementation of the sodium warning icon. Although decreases in purchases of high-sodium items among NYC FSR respondents were not significant relative to changes in purchases made by Yonkers respondents, both the mean sodium and calorie content of purchases made at NYC FSR declined significantly compared to Yonkers. This change may have been driven by the substantial reduction in the mean number of high-sodium items purchased by NYC FSR patrons. However, at baseline, FSR purchases of high-sodium items and mean sodium content were much higher in NYC than in Yonkers for unknown reasons. Although these analyses accounted for potentially confounding covariates such as demographic characteristics, which significantly differed between cities/years, and excluded extreme outliers, other factors such as location-specific promotions may have contributed to the baseline purchasing differences between NYC and Yonkers.

At QSR, there were no observed changes in high-sodium item purchases or in the sodium or calorie content of purchases in NYC compared to Yonkers. Assuming that the sodium warning icon did play a role in the reductions observed at NYC FSR, it is possible that a parallel impact was not seen in the QSR environment because of the differences in ordering method; at FSR, patrons presumably spend a longer time reviewing the menu book than QSR patrons may spend looking at the menu board.

While this study is, to our knowledge, the only real-world evaluation of consumer purchases following implementation of the NYC sodium warning regulation, two studies of hypothetical purchasing responses to sodium warning icons among online panels have been conducted. Byrd et al. tested whether hypothetical meal choices, when presented with sodium labeling, differed across taste preferences, with null findings for NYC’s sodium warning icon and mixed results for numeric sodium labeling [16]. Musicus et al. evaluated the effectiveness of various sodium warning icon designs, with positive findings for icons that included “Sodium Warning” text [17].

During data collection, NYC was the only jurisdiction in the United States with sodium warning labeling in the restaurant environment. Since then, Philadelphia implemented a similar sodium warning policy (their design informed by Musicus et al.’s research) in chain restaurants in 2019 [13],^13^ although no subsequent evaluations have been published.

The present study has several limitations. Findings cannot be generalized, as the sample was limited to patrons at 2 FSR and 2 QSR chains, and to those with an itemized receipt. Although participants indicated which items were for their personal consumption, it is unknown how much they consumed. While chain restaurants with consistent menus nationwide were chosen, it is possible that promotions and pricing differed across locations. Baseline and follow-up data were collected during different seasons, with baseline data collection occurring during fall and early winter, and follow-up during spring; although conducting a difference-in-differences analysis with a control city helps mitigate this limitation, changes from baseline to follow-up may have been influenced by the seasonal change. Finally, we were not adequately sampled to examine whether purchases differed by race, ethnicity, and other socio-demographic factors.

## Conclusions

While efforts like nutrient warnings aim to foster transparency about the food supply and may contribute to health-promoting behaviors [18], commercial factors such as excessive marketing, targeted marketing, and unhealthy food offerings can strongly influence health [19]. The accompanying menu evaluation by Sisti et al found no reduction in high-sodium offerings at chain restaurants approximately 18 months following implementation of the sodium warning icon; it may take additional time and action to reduce sodium in restaurant foods. Recently, the US Food and Drug Administration released voluntary sodium reduction goals for the food industry [20]. This, along with restaurant sodium labeling policies from additional jurisdictions, may prompt reformulation and lower-sodium offerings in the restaurant environment, which may have a larger effect on consumer purchasing. Further research on longer-term changes following such policies is warranted. Additionally, as new menu labeling policies are debated and passed (most recently, NYC’s Sweet Truth Act [21]), ongoing evaluation of label designs should be conducted to maximize the likelihood of their usefulness to consumers.

## Data Availability

Datafiles and code will be made available on GitHub following manuscript acceptance.

## Acknowledgements

The authors thank the following colleagues for their thoughtful contributions: Aldo Crossa and Sungwoo Lim for statistical guidance, Danielle Gurr, Elizabeth Kelman, and Lauren Shiman for equity-focused review, Shadi Chamany and Hannah Helmy for comments on the manuscript.

## Supporting Information

**S1 Fig: NYC chain restaurant sodium warning** icon (a), displayed next to all items containing 2,300 mg of sodium or more, and (b) sodium warning statement, posted in each restaurant at the point of order.

**S1 Table: High-sodium items*, IHOP, 2015 and 2017** *Included items that either: 1) displayed a warning icon in 2017 at the time of data collection, or 2) were not on the menu in 2017, but were ordered in 2015 and contained >= 2,300 mg sodium.

**S2 Table: High-sodium items*, TGI Friday’s, 2015 and 2017** *Included items that either: 1) displayed a warning icon in 2017 at the time of data collection, or 2) were not on the menu in 2017, but were ordered in 2015 and contained >= 2,300 mg sodium.

**S3 Table: High-sodium items*, Popeyes, 2015 and 2017** *included items that either: 1) displayed a warning icon in 2017 at the time of data collection, or 2) were not on the menu in 2017, but were ordered in 2015 and contained >= 2,300 mg sodium.

**S4 Table: High-sodium items*, Subway, 2015 and 2017** *included items that either: 1) displayed a warning icon in 2017 at the time of data collection, or 2) were not on the menu in 2017, but were ordered in 2015 and contained >= 2,300 mg sodium.

**S5 Table: Square root-transformed sodium and calorie content of purchases at full-service and quick-service restaurant chains, pre- (2015) and post- (2017) implementation of the sodium warning icon in NYC**

